# Stakeholder Perspectives on an Adult Cerebral Palsy Community Registry: A Qualitative Study

**DOI:** 10.1101/2024.09.26.24314382

**Authors:** Cristina A. Sarmiento, Edward Hurvitz, Jocelyn Cohen, Mary Gannotti

## Abstract

**Background:** The Cerebral Palsy Research Network (CPRN) community registry has yielded valuable information about changes in function and pain in adults with cerebral palsy (CP) through a patient-reported outcomes registry. However, it requires increased enrollment and diversity of participants to produce more generalizable conclusions.

**Objective:** To identify stakeholder perspectives about the barriers and facilitators to enrollment in the CPRN Community Registry, strategies to enhance recruitment efforts, and important questions for the registry.

**Methods:** Qualitative descriptive study using iterative focus groups, followed by inductive thematic analysis. Participants included adults with CP and caregivers, clinical investigators, and community leaders in the CP and disability spaces. We explored perspectives about motivations for registry participation, barriers and facilitators to participation, and strategies for increasing and enhancing diversity of enrollment.

**Results:** We conducted four focus groups (20 participants with lived experience; 10 clinical investigators; 9 community leaders). All participants valued the information provided by the registry and felt that ongoing data collection was important. Barriers and related facilitators to participation include benefits of participation, awareness, accessibility, and collaboration with community and clinical partners. Adults with lived experience seek more precisely defined health and function outcomes for adults with CP.

**Conclusions:** Adults with lived experience, clinical investigators, and community leaders identified barriers and facilitators to participation in a patient-reported registry and important questions. Our study revealed that communicating a direct benefit to the participant, improved visibility and accessibility, leveraging collaboration with clinical and community partners and answering more precise research questions could promote enrollment.

## Introduction

By systematically collecting and analyzing patient-reported data, a community-based registry can be a powerful tool to observe the course of a condition, understand variations in treatment and outcomes, and examine factors that influence prognosis and quality of life.^1^ This type of information is critical for policy, clinical practice, and program planning. The need for such a registry to accelerate practice and research around aging and cerebral palsy (CP) led to the development of the Community Registry Adult Surveys on Function & Pain by the Cerebral Palsy Research Network (CPRN).^2^ Although the initial reports from the registry provided important information about antecedents for functional decline and chronic pain characteristics and treatments, the low enrollment and lack of diversity among respondents limits generalizability of these findings.^3,4^ Engagement of stakeholders (i.e., individuals with lived experience, clinicians, and community organizations), is needed to identify facilitators and barriers to enrollment in the registry, and to develop effective recruitment strategies for diverse communities.

The CPRN is a learning health network for CP consisting of more than 30 clinical sites and bringing together clinicians, researchers, and persons with lived experience (i.e., individuals with CP, family members, caregivers) to improve health outcomes for people with CP.^5^ The number of clinical sites involved in the network and the number of community organizations affiliated with CPRN results in large reach to caregivers and individuals with CP. Additionally, CPRN has large community engagement through a web-based platform, MyCP. Leveraging this large infrastructure more effectively could increase enrollment in the registry substantially.

The CPRN Community Registry and its associated surveys were co-created by consumers, clinicians, and researchers to improve the quality of life and care for adults with CP.^2^ Reflecting on the relatively low enrollment with lack of diversity, collaboration with stakeholders is needed to identify effective, feasible solutions.^6^ Currently enrollment in the registry is passive, and members of CPRN’s MyCP community who meet eligibility criteria have the opportunity to participate through CPRN’s MyCP platform.^2^ Demographics of the initial enrollees include predominantly college educated, White women who ambulate.^3,4^ College-educated women can be more likely to answer internet surveys at large.^7^ Enrolling Black, Indigenous, and People of Color will take intentional recruitment strategies,^8,9^ as will enrolling people with more extensive functional limitations.^10^

Engaging individuals with lived experience in all stages of the research process facilitates the production of knowledge that is relevant and important to those with lived experience, including individuals with lifespan disabilities.^11-13^ Furthermore, intentionally co-creating solutions with stakeholders is an effective way to create research that can lead to meaningful social change.^6^ Engaging clinicians and community organizations serving adults with CP will facilitate additional insights and recruitment strategies, given the unique roles these collaborators play in the lives of individuals with CP. The objective of this study was to identify barriers and facilitators to registry enrollment, potential recruitment strategies, and identify important questions to be answered by the registry by engaging individuals with lived experience, clinical investigators, and community leaders.

## Materials and Methods

### Study Design

We conducted a qualitative descriptive study using iterative focus groups to gather input from people with lived experience, community organizations, clinicians, and researchers regarding barriers and facilitators to registry enrollment, recruitment strategies, and important questions to address using registry data. Our study was exempted by the Colorado Multiple Institutional Review Board (Protocol #23-1653). We report our methods and findings using the Standards for Reporting Qualitative Research.^14^

### Participants and Procedures

We conducted three rounds of focus groups beginning with people with lived experience, followed by clinical investigators, and then community organizations and leaders. Each focus group began with knowledge sharing and education related to the identified need for the registry, its development, initial results, and challenges with enrollment. This was followed by participant input on barriers and facilitators to participation using a semi-structured interview guide (See Appendix 1). After each round of focus groups, thematic analysis and exemplar quotes were integrated into the presentation for the subsequent group to build iteratively on previously collected data.

Hence, the community leaders were presented with data reflecting the perspectives of the people with lived experience and the clinical investigators. The iterative nature allowed for subsequent focus groups to build upon each other. Having separate focus groups for people with lived experience, clinical investigators, and community leaders created an environment that encouraged free sharing of opinions, and active listening, response, and experience sharing between participants.

Adults with CP and caregivers were recruited from study team clinical sites, CPRN, and Cerebral Palsy Research Alliance Foundation (CPARF) stakeholder groups. Inclusion criteria for adults with CP were: 1) 30 years of age or older and 2) a self-reported diagnosis of CP. Caregivers were those who self-reported that they provide care for an adult with CP. Enrolling adults with CP 30 years or older allowed time for people to have more lived experience as an adult with CP and may have begun to consider impact of aging on function and pain. We purposefully sampled for diversity of gender, race and ethnicity, and functional mobility level, as these characteristics may also influence experiences with barriers and facilitators to participation in surveys. We aimed for two focus groups of individuals with lived experience, consisting of 8-10 participants each, with separate groups based on functional mobility level (i.e., primarily ambulatory and primarily non-ambulatory).

Clinical investigators were investigators from CPRN sites and other CP-focused clinical sites, who self-identified as regularly providing care to adults with CP or involved in the transition from pediatric to adult care. They were also investigators interested in multi-site research or using the learning health network of CPRN as a way to advance care for adults with CP. We purposefully sampled for diversity in clinical role (e.g. MD, PT) and specialty (e.g., neurology, physical medicine and rehabilitation). We planned for one focus group of clinicians with 8-10 participants.

Community leaders and disability advocates were recruited from CP and disability community organizations throughout the nation as well as social media advocates in the CP and disability spaces. We purposefully sampled for diversity in types of organizations and platforms. We planned for one focus group of community leaders with 8-10 participants.

Using maximum variation sampling and snowball sampling, both types of purposeful sampling,^15-17^ we recruited focus group participants for diversity of participant background based on characteristics detailed above. In maximum variation sampling, the research team identifies key variables and then identifies cases that vary from each other as much as possible, allowing the research team to identify meaningful patterns from a diverse sample.^17^ In snowball sampling, the study team seeks information about other information-rich cases or participants from key informants themselves. ^17^

We developed the focus group guides (Appendix 1), based on our study’s objectives, prior research and experience using CPRN Community Registry data, and input from CPRN clinicians and community members during webinars detailing this study’s proposed methods. Focus groups were conducted using secure videoconferencing (Zoom; San Jose, CA), and lasted approximately 90 minutes. A non-clinician member of the study team who is an adult with CP (JC) facilitated focus groups to minimize the potential bias of having a clinician facilitator. We obtained verbal consent for participation and recording from each participant at the start of each focus group. Within each group, the study team first shared knowledge about the background for creating a patient-reported registry, its development, the initial findings, and its current challenges with enrollment. Then, the facilitator guided conversation to further explore participant perspectives on important questions to be answered by the registry, potential barriers and facilitators to enrollment, and recruitment strategies to enhance diversity of enrollment. During focus groups with clinical investigators and community leaders, we also assessed perceived importance of having a clinical registry via Zoom polling function. We audio-recorded focus groups with permission from participants, and recordings were professionally transcribed (Landmark Associates; Phoenix, AZ). Transcripts were de-identified prior to analysis. Ahead of focus groups, participants also provided self-reported demographic data into a secure REDCap electronic database hosted at the University of Colorado Anschutz.^18^

### Data Analysis

Using a qualitative descriptive approach to analyze data, we sought to explore, understand, and describe participants’ varied perspectives.^19-21^ Three members of the study team (CS, MG, EH) used an inductive approach to coding transcripts by creating codes based on emergent elements in the data.^22^ To develop a final codebook, we met regularly to reconcile and calibrate coded transcripts.^23^ At least two of the three coding team members double coded all transcripts. We then categorized and grouped together responses and dialogue of the participants in the focus groups. We reviewed and summarized quotations and identified salient themes using thematic analysis.^23,24^

### Results

In total, we conducted four focus groups (20 participants with lived experience; 10 clinical investigators; 9 community leaders). Table 1 provides demographic information on the participants with lived experience: 25% were non-White, 45% male, mean age 49.6 years, with a distribution of gross motor abilities from independent to dependent and from various community settings, including rural settings. Figure 1 displays the geographic diversity of participants in all the focus groups. Forty-two percent of all participants were from the Northeast and 24% were from the Midwest, with the remaining participants from the Southwest (11%) , Southeast (13%), and West (11%). Most all the participants were from suburban (47%) and urban (44%) areas as compared to rural (8%) areas. Figure 2 displays the variety of specialties of participating clinical investigators and variety of populations served by the community organizations.

**Table 1.** Characteristics of adults with cerebral palsy and caregivers participating in focus groups (N=20).

| **Self-Reported Characteristics** | **n (%) unless otherwise specified** | **Missing** |
| --- | --- | --- |
| **Role** |  | 0 |
| Adult with CP | 19 (95%) |  |
| Caregiver | 1 (5%) |  |
| **Age (years):** Mean, SD (range) | 49.6, 12.8 (33-72) | 1 |
| **Female Gender** | 11 (55%) | 0 |
| **Race** |  | 0 |
| Asian | 2 (10%) |  |
| Black or African American | 2 (10%) |  |
| White | 15 (75%) |  |
| Choose not to answer | 1 (5%) |  |
| **Ethnicity** |  | 0 |
| Hispanic | 1 (5%) |  |
| Not Hispanic | 18 (90%) |  |
| Choose not to answer | 1 (5%) |  |
| **GMFCS level** |  | 1 |
| I | 4 (20%) |  |
| II | 6 (30%) |  |
| III | 5 (25%) |  |
| IV | 3 (15%) |  |
| V | 1 (5%) |  |
| **Mobility Function** |  | 3 |
| Walks independently always | 6 (30%) |  |
| Walks independently sometimes | 1 (5%) |  |
| Walks with assistance always | 2 (10%) |  |
| Walks with assistance sometimes | 6 (30%) |  |
| Walks never | 2 (10%) |  |

*CP – cerebral palsy*
*GMFCS – Gross Motor Function Classification System*

**Figure 1.**
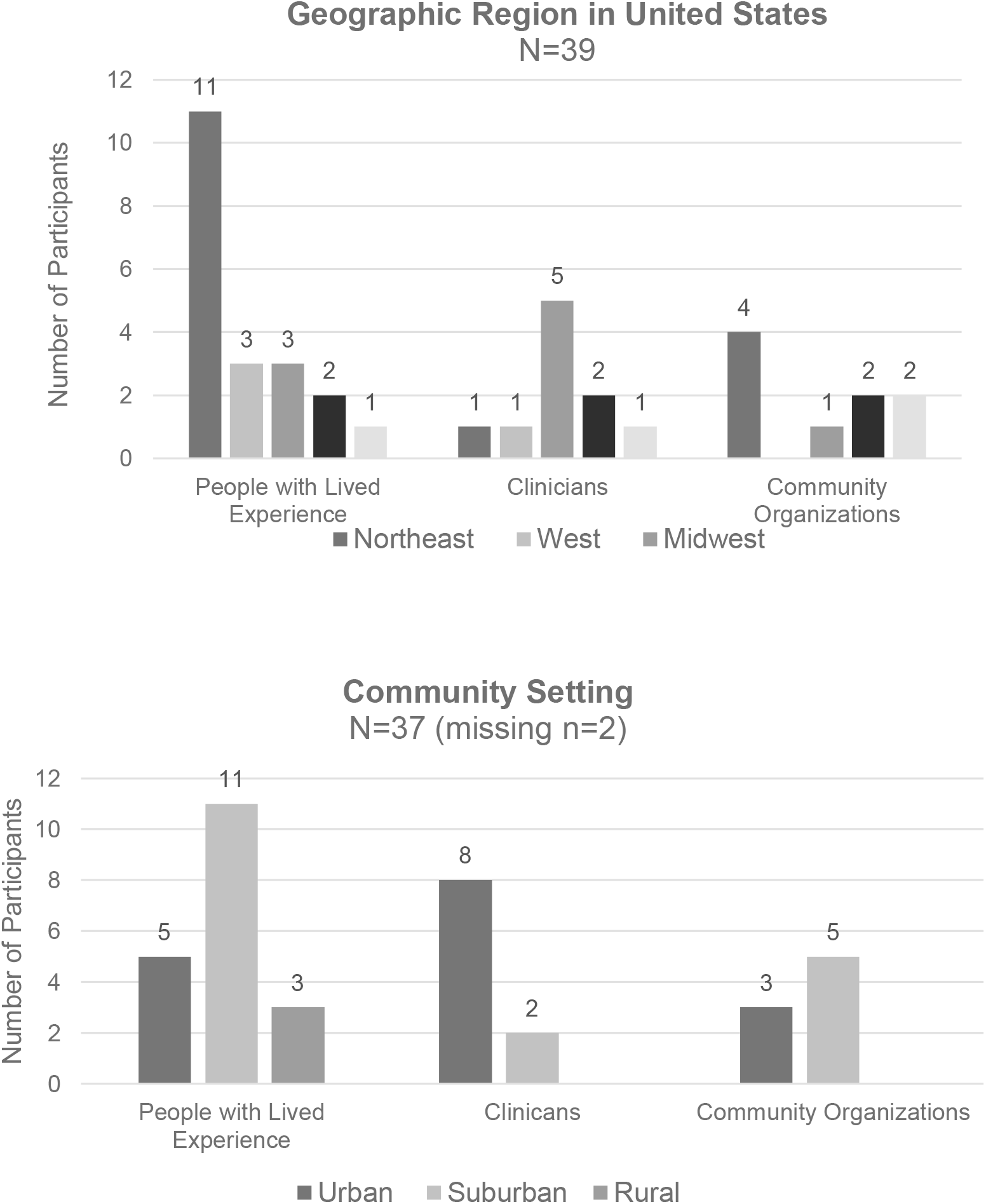
The geographic diversity of participants in all focus groups by subgroups.

**Figure 2.**
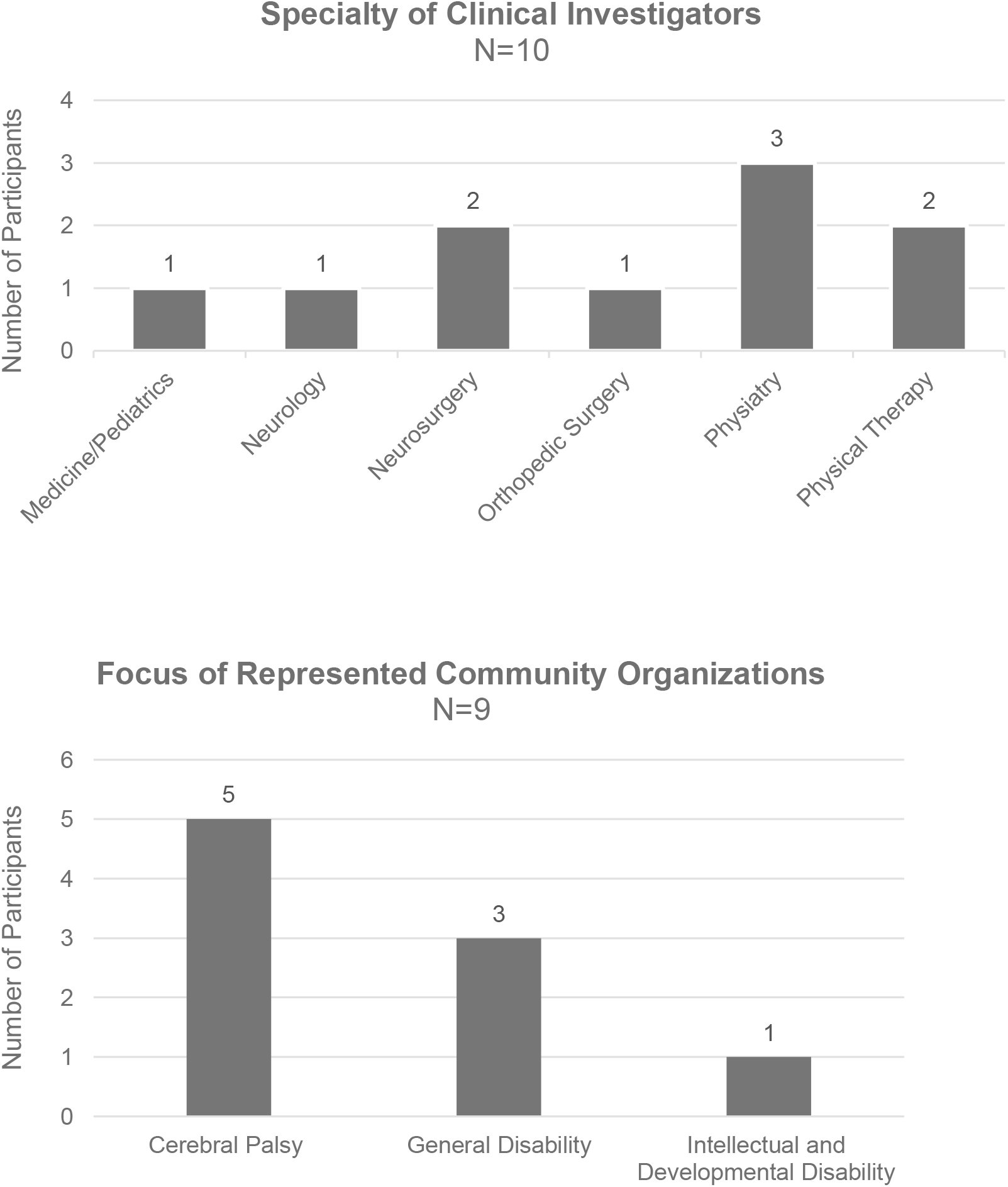
The variety of specialties of the participating clinical investigators and the populations served by community organization leaders.

Participants were asked about the importance of having a patient-reported registry for adults with CP, and generally felt that this was very important. We identified two themes regarding participation: the importance of appropriately communicating motivators and ensuring accessibility. Table 2 presents each theme with codes, barriers, facilitators, and solutions. Identified recruitment strategies focused on maximizing opportunities with collaborators and leveraging community connections. Research questions important to participants included questions about individualized outcomes or “precision rehabilitation”. Solutions were proposed by participants for all current barriers and for increasing collaboration with clinical settings and community organizations. Table 3 presents codes related to recruitment strategies and research questions along with participant-identified barriers, facilitators, and solutions.

**Table 2.** Identified themes and codes, and participant-identified barriers, facilitators, and solutions.

| Theme | Codes | Barriers | Facilitators | Solutions |
| --- | --- | --- | --- | --- |
| **Appropriately Communicating Motivators** | Self-Knowledge | Questions not specific enough<br>Registry not designed to provide personalized recommendations | Want to know more about one's health<br>Results of survey are given to participant with ratings compared to others | Continue to provide individual reports to those who participate |
|  | Direct Benefit | Registry not designed to directly benefit an individual | Communicate participant- identified benefits (learn more about one's experiences, contribute to community knowledge) | Provide more tangible reason for participation, more clearly communicate these benefits during recruitment |
|  | Trust | N/A | CPRN viewed as reputable source | Continue to leverage CPRN infrastructure and connections |
|  | Contribute | Survey burnout | Help increase knowledge about cerebral palsy and aging | Have specific questions about specific conditions important to participants<br>More clearly communicate benefit to broader community |
| **Ensuring Accessibility** | Lack of Awareness | No awareness of the registry | Webinars<br>CPRN social media presence | Visual advertisements<br>Infographics for advertisement |
|  | Universal Design | Not enabled for ease of use with eye gaze, head pointer, or other adaptive interfaces.<br>Registry not accessible to people with cognitive or other executive function disorders | Ability to stop survey and then come back to finish without losing answers | Explore the universal design interface for survey to ensure accessibility<br>Explore caregiver report registry for adults who have intellectual disability |
|  | Ease of Use | Multiple layers of sign in before access surveys | Streamline access, minimize number of steps | Modifications to application to reduce user burden |

*CPRN – Cerebral Palsy Research Network*

**Table 3.** Participant-identified recruitment strategies and research questions.

| Topic | Codes | Barriers | Facilitators | Solutions |
| --- | --- | --- | --- | --- |
| **Recruitment Strategies** | Minimize Burden | Survey burnout, length of registry surveys | Aggressive intentional retention strategies to optimize survey completion<br>Allow breaks without losing answers | Use of modules to split up surveys<br>Reduce the number of surveys<br>Prioritize surveys important to participant-identified questions |
|  | Maximize Opportunities | Institutional agreements and funding/time | Distribution of information about the registry in clinic and hospital settings | Use of QR code directing to MyCP for sign up<br>Obtain permissions for active recruitment in clinic |
|  |  | Organization agreements and funding/time | Opportunities with organizations, existing connections and collaborations | Collaboration with information dissemination to communities served<br>Active recruitment in broader disability spaces |
|  | Diversity | Missing diverse population | Broader disability and community space | Outreach with specific sports clubs and at disability specific sites |
| **Research Questions** | Precision Medicine | Current questions are broad as there is limited information on adults with cerebral palsy | Desire for information about outcomes for people with particular characteristics: age, gender, gross motor ability, life course. | Modify research questions, continue to co-create with the consumers<br>Create specific research questions to address these questions |

### Appropriately Communicating Motivators

Motivators for participation in the registry identified by consumers were: educating oneself on issues important to aging with CP, contributing to increased scientific knowledge, and trust in CPRN as a reputable source for information and research opportunities. Not understanding the direct benefit was a deterrent to participation. Those participants that had participated in the CPRN Community Registry often cited the desire to help advance knowledge and give back to their community as motivating factors. They hoped that sharing their story might help improve future care, for themselves and other adults with CP to come.

> *I took it. It was basically twofold. Because, when things started happenin’ that I wasn’t prepared for, I needed answers, and I wasn’t stoppin’ until someone [laughs] told me what the answers were. When I started searchin’ for the answers, that’s when I discovered there weren’t many to be had. Then I connected with [a CPRN investigator], I think, when CPRN first started. I’ve just been in it the whole time just for the answers and to try to spread awareness so no one would ever have to go through the ordeal that I went through because the answers weren’t there*. (Participant 7, adult with CP)

Other participants also felt that participating in the Community Registry helped them understand their own health, and that they were not alone with the issues they were facing in adulthood.

> *When I was growing up with CP, obviously, there was nothing much out there, ‘cause I’m [in my 70s] right now. Over the years, I kinda was in denial that I had CP…I was in denial, and then I started realizin’ that I had a problem. I didn’t understand why. I got ahold of one doctor, and he gave me medication for stuff, but I didn’t really understand CP and how it really affected me. Then I started bein’ more aware. The doctor was gonna retire, and he gave me the connection to CPRN. I started to do all these surveys and all the questions. It was a learning experience because every question they had, I says, “Oh, that’s what’s going on. Oh, wow, I never realized this*.*” It helped me understand what I was dealin’ with with CP*. (Participant 5, adult with CP)

Relatedly, one participant felt that it was challenging to find “reputable” sources of information on aging with CP, and the Community Registry as well as CPRN helped to provide some of this:

> *I think, as a person with CP, I would like to be more informed. There needs to be more information channels for us to—more reputable information channels that we can access to provide information on the aging process with people with cerebral palsy and what we can expect…It would be nice to have a resource I could go to, other than my primary care doctor, where I could learn more about what’s going on as I age, as a person, particularly a woman, with CP*. (Participant 1, adult with CP)

One participant emphasized the need to clearly convey *why* they should participate and felt this would help motivate others to participate, too:

> *Because a lotta people get asked to fill out a lotta surveys and participate in a lotta different groups and volunteer information and time. The “What’s in it for me?” I think is really valuable. People are motivated, obviously, as you all well know, by very different things. Some people are motivated just by wanting to be of service and helpful and useful. Other people, really, are like, “Hey, what am I gonna get out of this? What’s the concrete benefit?”* (Participant 13, adult with CP)

### Ensuring Accessibility

Lack of accessibility of the registry, both general public awareness about its existence and its ease of use by a variety of users, were cited as barriers to enrollment. To increase general public awareness, participants suggested using more visual information, infographics, and social media presence to promote the registry. Participants with lived experience and community organization members cited difficulty in accessing the surveys, finding out about them, signing up for MyCP, and then logging in to the survey portal as barriers to completing the registry surveys.

Additionally, several community leaders noted how the survey did not include web browser accessibility features to support individuals who used technology such as head pointers or eye gaze to activate the computer.

> *I’m thinking of adults with CP who maybe rely on a caregiver. They’re relying on that caregiver’s skill in order to create an account and then access a survey and that brings in a lot of questions around technology usage and then just about the accessibility as well*. (Participant 37, community leader)

The surveys also do not provide accommodations for people with poor self-regulation (e.g., needing to take breaks during the survey) or intellectual disability. Currently, the Community Registry only collects data from adults with CP who can consent and complete the surveys themselves and does not allow for proxy responses or caregiver consent/participation. One participant with lived experience emphasized the importance of collecting registry data about adults with CP who cannot consent themselves:

> *CP’s a very large and diverse spectrum, and not everybody is able to verbally communicate and/or have gone to post-secondary in some capacity. I think it’s very important to keep that in mind*. (Participant 12, adult with CP)

### Recruitment Strategies

Participants suggested multiple possible recruitment strategies to address current enrollment limitations. Recruitment could be optimized by minimizing respondent burden and leveraging opportunities with community and clinical collaborators. In addition to the accessibility of the surveys as mentioned above, there was a concern about the number of survey questions asked. One participant noted.

> *“… [we are working] pretty aggressively [to] reduce barriers. We saw that a lot of our own folks locally who were getting a few questions in or partially through a survey and then just not completing them⃛ we’ve ended up making them more modular, making them shorter, making various options for shorter surveys*.*”* (Participant 25, clinical investigator)

Participants identified recruitment opportunities in collaborating with CPRN-affiliated clinical sites and community organizations. Participants felt that direct recruitment through CPRN as well as recruitment through social media would be effective strategies. Participants suggested having CPRN send out more tweets about the Community Registry and utilize more infographics to convey importance of participation.

Clinical investigators suggested more active recruitment efforts in clinical spaces (e.g., flyers in clinic waiting rooms, research assistants at clinical sites directly contacting and enrolling participants). Currently, however, recruitment into the registry is passive, and people can only be directed to MyCP (i.e., rather than being enrolled into the registry directly). Still, opportunities were identified by the clinicians for increasing awareness about the registry by using QR codes that link to MyCP in discharge notes, visit summaries, and flyers in clinic waiting rooms.

Community collaborations suggested by participants included organizations such as: Centers for Independent Living, University Centers of Excellence for Developmental Disabilities, the Department of Rehabilitation Services, Vocational Rehabilitation, or at disability expos or events, which would target the disability space more broadly. Additionally, recruitment through universities or high schools could help recruit more older adolescents and young adults, while recruitment through community centers and senior centers, could help target older adults. Supportive care or independent care facilities may help recruit more individuals with severe mobility impairments and/or wheelchair users. Adaptive sports programs and recreation centers may help recruit more active and possibly more male participants. Other ideas were to hold information sessions to target adults with CP in the community by educating them about the importance of and benefits from participating in the registry.

### Important Research Questions

Participants with lived experience identified many questions that they would be interested in answering using Community Registry data. Of particular interest was how differences in CP type, gender, and life experiences impact outcomes. Some of the participant-identified questions included:

- What are the unique experiences of women with CP (gynecologic health, pregnancy, childbirth, etc.)? How do these affect pain, tone, and function?
- Among adults with CP, how do different genders vary – particularly in terms of pain?
- How can the Community Registry be expanded/adapted to collect information on adults with CP by caregiver proxy?
- What can be done to prevent pain as adults with CP age? (therapy and exercise protocols, equipment, positioning)
- How can workplace accommodations help minimize pain and functional decline?
- How does pain differ for adults with CP?

∘ How do different types of pain relate to/co-occur with each other?
∘ How do management strategies differ?

- What types of exercise are adults with CP doing? What unique benefits and challenges exist for different types, and how do people manage the challenges?

Many participants prioritized research related to pain, and multiple adult women with CP expressed that the Community Registry could help us better understand women’s health as it relates to CP and pain. As one participant shared:

> *I was thinking about looking into some of the gender differences, ‘cause they—I think there’s been studies saying that females sometimes can experience more pain. I would be curious to know why…I think we definitely need just more research in general on health, but particularly female health because female is different from males in different ways*. (Participant 12, adult with CP)

Additionally, participants with lived experience were looking for information about specific issues that were a combination of multiple factors (e.g., age, gender) in addition to cerebral palsy. Clinicians and community organization leaders were also interested in linking specific clinical characteristics, personal characteristics, treatment and outcomes across the lifespan in a personalized medicine approach.

## Discussion

We sought to explore stakeholder perspectives about barriers and facilitators to enrollment in the CPRN Community Registry, potential recruitment strategies, and important questions to be answered by the registry. We conducted iterative focus groups with adults with CP, caregivers, clinical investigators, and community leaders using a qualitative descriptive approach to data analysis. Participants identified issues with awareness and accessibility of the registry, the importance of appropriately communicating the importance of the registry, opportunities for collaboration with community and clinical partners, and important questions to more precisely define health and function outcomes for adults with CP. Next steps include incorporating identified recruitment strategies, refining questions to be answered by the registry, and leveraging opportunities to increase enrollment.

To meet goals of increasing the number and diversity of enrollees in the Community Registry Adult Surveys on Function and Pain, solutions generated were similar to strategies developed by other health researchers.^11,12^ Motivators to participate included perception of a direct benefit and obtaining answers to more specific questions, in addition to the opportunity to share one’s experience to help the collective population of people with CP. Collecting data on issues important to community stakeholders could help increase motivation to participate in the registry. Other researchers also recommend co-creating questions for similar studies by identifying a specific need within the community to increase inclusion of people with disabilities in research.^11,12^

Issues related to access have been cited by others as the primary barrier to participation for minorities and people with disabilities.^13,25^ Black and Hispanic individuals are as likely or more likely to participate and follow up in longitudinal studies as White individuals, though access is a primary barrier.^25,26^ We also identified access as a theme, both access related to knowledge of the registry’s existence and accessible interfaces to support enrollees with different abilities. To increase representation of persons with disabilities in research, designing the research so most people can participate without the need for adaptions, or to allow for the use of one’s adaptations, was recommend by participants, and it has been suggested by other authors as well.^13^ Both proactive (face-to-face) and reactive recruitment strategies (collaboration with key leaders, snowball and word of mouth information sharing, printed material, broadcast media) as well as being flexible, building rapport and trust, and employing research staff with diversity in race, ethnicity, and ability are strategies that have been identified as effective.^27^

The CPRN Community registry has potential to provide insight into knowledge gaps about precision rehabilitation for adults with CP. Precision rehabilitation is a patient-centered approach to rehabilitation that aims to maximize a patient’s function and minimize disability by providing the right therapy at the right time.^28^ It is based on the idea that linking biology to behavior can help break unhealthy cycles and improve quality of life.^29^ People with lived experience with CP are seeking these types of answers for optimizing quality of life with aging, as shown by prior studies and the research questions identified by participants in this study.^30^ Clinicians, researchers, and people with lived experience with other neurologic disabilities are seeking similar answers.^31,32^

### Study Limitations

Our study has several limitations. First, participants were recruited from CP- and disability-focused networks. As such, findings may not be transferable to the general community, specifically to people who are less connected with and less active in CP research and advocacy efforts.^33^ Further, the majority of lived experience participants were White and non-Hispanic, though we successfully recruited several participants with lived experience who were non-White and/or Hispanic. Second, while it is important and was our priority to amplify the voices of adults with CP in research, our lived experience focus groups only included one caregiver; therefore, the perspectives of a larger group of caregivers may differ.

## Conclusions

A longitudinal, patient-reported outcomes registry that tracks changes in function and pain over time can have an important impact on clinical care, policy, and programming for adults with CP. The CPRN Community Registry Adult Surveys on Function and Pain have, to date, provided cross-sectional information that has contributed to our knowledge about adults with CP and aging. Increased enrollment with more diverse participants will provide both more precise and generalizable information. The information gleaned from this project provides solutions and strategies to advance and optimize the recruitment and enrollment of community members in the registry.

## Data Availability

All data produced in the present study are available upon reasonable request to the authors.

## Acknowledgements

We would like to thank the adults with cerebral palsy, caregivers, clinical investigators, and community leaders who participated in our study, sharing their experiences and perspectives with the hopes of improving health, function, and quality of life for individuals with cerebral palsy across the lifespan.

## Notes

### Competing Interest Statement

Jocelyn Cohen is affiliated with the Cerebral Palsy Alliance Research Foundation (CPARF). However, she did not participate in grant review or funding decisions. The grant and funding committees of CPARF did not participate in the study design, data collection or analysis, or manuscript preparation. No other competing interests to declare.

### Funding Statement

This work was supported by the Cerebral Palsy Alliance Research Foundation (CPARF) who funded an Accelerator Award for work within the Cerebral Palsy Research Network. The funder had no role in determining the funding, design, implementation or interpretation of the findings.

### Author Declarations

The Colorado Multiple Institutional Review Board deemed this work exempt (Protocol #23-1653).

