## Appendix 1 for "Stakeholder Perspectives on an Adult Cerebral Palsy Community Registry: A Qualitative Study"

**Appendix 1. Focus Group Guides**

**People with Lived Experience**

1. If you have participated in the CPRN’s Community Registry, what encouraged you to participate?
   a. If you have not participated, what might encourage you to participate?
      i. [Probe, if needed] What concerns or difficulties do you have about participating?
      ii. [Probe, if needed] What could be better?
   b. What would be an effective way to communicate opportunities like this?
      i. [Probe, if needed] What sources do you go to for information like this?
2. What are some ways to help encourage adults with CP to participate in research?
3. What are the questions that you would like answered about how function and pain changes and is treated?
4. We reviewed the characteristics of current participants in the Community Registry. How can we encourage more diverse adults with CP to participate (e.g., males, wheelchair users, and persons with diverse racial and ethnic backgrounds)?
5. What questions and/or actions are most important to address first from the findings we discussed?

**Clinical Investigators**

1. What supports are needed for adults with cerebral palsy within your clinic to be enrolled in MyCP, prior, during or within a clinic visit?
2. What unique features exist in your clinic, community, region, that may facilitate sharing of information about the CP Network and the Adult Surveys?
3. Which of these strategies seem most effective? Why?
4. Which of these strategies seem most feasible? Why?
5. How important is it to collect prospective longitudinal patient reported data from adults with cerebral palsy living in the community on a scale of 1 to 5 with 1 being least important and 5 most important?

**Community Organization Leaders**

1. Which of these strategies seem most feasible? Why?
2. Which of these strategies seem least feasible? Why?
3. Which of these strategies seem most effective? Why?
4. Which of these strategies seem least effective? Why?
5. How do you think that your organization might be able to engage in these recruitment strategies?
6. How important is it to collect prospective longitudinal patient reported data from adults with cerebral palsy living in the community? Rate this on a scale of 1 to 5 with 1 being least important and 5 most important. Please answer via the Zoom poll.
7. What unique features exist in your organization that may facilitate sharing of information about CPRN and the Community Registry?
